# Volumetric Integrity of Frontotemporal and Perisylvian Regions for Predicting Behavioral and Language Impairment in Behavioral FTD: A Structural MRI Study

**DOI:** 10.64898/2026.09.30.26364390

**Authors:** Amir Hesam Zare, Iman Kiani, Leila Zeinalzadeh, Aydin Taghilou, Zahra Mirsepasi, Vajiheh Aghamollaii

## Abstract

**Background:** Behavioral variant frontotemporal dementia (bvFTD) is clinically defined by progressive changes in personality, executive function, and language, yet the structural neural correlates of symptom severity remain unclear. This study aimed to assess the association between clinical findings and neuroimaging biomarkers.

**Methods:** We included 87 patients with bvFTD. Clinical functioning was covering behavior and language. Spearman’s rank correlations were used to assess univariate associations, and Bonferroni correction was applied for multiple comparisons. Principal component analysis (PCA) was conducted on regional volumes, and multivariable logistic regression was used to model high behavioral impairment (FBI total ≥30), adjusting for age, sex, and education.

**Results:** No individual regional volume was significantly correlated with clinical scores after Bonferroni correction (all adjusted p>0.05), and PCA-derived imaging components failed to predict behavioral severity in multivariable models. Age emerged as the only significant predictor (OR=1.11 per year; p=0.009). Model discrimination was modest (AUC=0.735) and did not improve with the inclusion of imaging features (p=0.453 vs. demographics-only model).

**Conclusions:** In this bvFTD cohort, structural volumes of key frontotemporal and perisylvian regions did not predict behavioral or language impairments after correction for multiple comparisons. These findings show the limitations of regional volumetric analysis in isolation and stress the need for network-based, multimodal, and longitudinal approaches.

## Introduction

Frontotemporal dementia (FTD) encompasses a group of clinically and pathologically heterogeneous neurodegenerative syndromes that primarily affect the frontal and temporal lobes [1]. It is a common cause of early-onset dementia [2] and is typically characterized by insidious changes in behavior, personality, and language [3]. Clinically, FTD can be divided into three primary variants: the behavioral variant (bvFTD), and two language variants, semantic variant primary progressive aphasia (svPPA) and nonfluent/agrammatic variant PPA (nfvPPA) [4]. These clinical phenotypes are underpinned by distinct patterns of brain atrophy, which can be identified and quantified using structural magnetic resonance imaging (MRI).

Among the brain regions consistently implicated in FTD, five structures stand out for their critical involvement in both behavioral regulation and language processing: the inferior frontal gyrus (IFG), pars opercularis (a subdivision of the IFG), insula, superior temporal gyrus (STG), and inferior parietal lobule (IPL) [5]. These regions are components of broader frontotemporal and perisylvian networks that support high-order executive, affective, and linguistic functions [6].

The IFG, particularly its left hemisphere, is a cornerstone of the language production network. It encompasses Broca’s area and is heavily involved in syntactic processing [7]. The pars opercularis, forming the posterior segment of the IFG, specifically supports phonological processing and articulatory planning [8]. The insula, a deeply situated cortical region, acts as a multimodal integration hub. It bridges sensory input, autonomic regulation, emotional salience, and interoceptive awareness [9]. The STG is a key node of the auditory and language comprehension network. Beyond language, this region supports social cognition, particularly theory of mind [10]. The IPL, though more traditionally associated with Alzheimer’s disease, has garnered attention in FTD for its role in complex language tasks and visuospatial processing. It is involved in semantic integration, phonological working memory, and sentence-level comprehension [11].

Given the multifocal and progressive nature of FTD, understanding the relationship between regional brain atrophy and specific clinical symptoms is crucial for both diagnosis and monitoring. Structural MRI provides a non-invasive method to quantify regional volumes and track neurodegeneration over time. Numerous studies have reported associations between focal atrophy and distinct behavioral or linguistic deficits in FTD. However, findings are often fragmented, limited to small samples, or focused on single clinical or anatomical features. A comprehensive approach that examines multiple brain regions across multiple clinical domains, while accounting for covariates such as age, sex, and education, is needed to clarify the neuroanatomical basis of functional decline in FTD.

In this study, we examined the volumetric integrity of five key brain regions, the inferior frontal gyrus, pars opercularis, insula, superior temporal gyrus, and inferior parietal lobule, and explored their associations with domain-specific clinical scores in FTD. We further applied multivariate modeling techniques to determine whether imaging-derived components could predict the presence of significant behavioral impairment, and compared their predictive value to that of demographic variables alone. Finally, we assessed whether sex-related anatomical differences influenced imaging measures or clinical performance. Through this integrative framework, we aimed to better understand the structural basis of language and behavioral dysfunction in frontotemporal dementia.

## Methods

The study was approved by the ethics committee of Tehran University of Medical Sciences and adhered to the principles outlined in the Declaration of Helsinki. All participants or their legal representatives provided written informed consent in accordance with institutional ethics guidelines. Participants were eligible for inclusion if they had complete baseline imaging data, as well as complete clinical scores and demographic information, including age, sex, and years of education. Records with missing values in any of these key analytic variables were excluded: pairwise deletion was applied in univariate analyses, while listwise deletion was used for multivariable models. All continuous variables were standardized (z-scored) prior to principal component analysis and regression modeling.

To explore bivariate relationships between regional brain volumes and clinical functioning, we calculated Spearman’s rank correlation coefficients (ρ) across all combinations of the five anatomical volumes and nine domain-specific clinical scores. Non-normality in several variable distributions was confirmed via Shapiro–Wilk tests. Bonferroni correction was applied by multiplying raw p-values by 45 and capping the results at 1.0. Statistical significance was set at an adjusted p < 0.05. Correlation results were visualized using heatmaps for both effect size (ρ) and adjusted p-values.

Principal component analysis (PCA) was conducted on the five z-scored regional volumes using the scikit-learn library in Python. Components with eigenvalues greater than 1.0 were extracted. Loadings were examined to interpret each component’s anatomical relevance.

We defined a binary outcome variable of high behavioral impairment, using a threshold of ≥30 on the total Frontal Behavioral Inventory (FBI). A multivariable logistic regression model was then fitted using the Logit function in the statsmodels package. Convergence status and coefficient magnitude (|β| < 10) were checked to rule out separation issues. Linearity of continuous predictors in the logit was evaluated by plotting binned means against log-odds. Influential observations were flagged using Cook’s distance with a threshold of >4/(n – k – 1). Calibration was evaluated using a 10-bin observed-versus-predicted probability plot. Model fit was assessed via the likelihood-ratio (LR) test comparing the full model to an intercept-only null model.

Model discrimination was quantified using the area under the receiver operating characteristic curve (AUC), and performance was summarized at the Youden’s J index. Model comparisons were based on AUC differences, significance testing via bootstrap resampling (n = 1000), and classification performance at the optimal threshold.

Sex differences were explored in two domains: anatomical and clinical. First, we assessed whether regional volumes differed by sex. After testing for normality (Shapiro–Wilk) and equality of variances (Levene’s test), we applied either Student’s t-test (equal variances), Welch’s t-test (unequal variances), or Mann–Whitney U test (non-normal distribution). Bonferroni correction was applied for five comparisons (α = 0.01). Second, each clinical score was dichotomized at its median, and sex differences in high vs. low scores were tested using Pearson’s χ^2^ on 2×2 contingency tables.

## Results

A total of 87 participants were included in the final analytic sample. The mean age was 63.6 years (SD = 6.7, range = 43–78), with a near-equal distribution of sex (48% male) and education status (52% with higher education). Behavioral assessments revealed a wide range of symptom severity: the mean total score on the Frontal Behavioral Inventory (FBI) was 34.7 (SD = 12.0), with its subscales, negative behaviors (FBI-NBS) and disinhibited behaviors (FBI-DBS), averaging 17.1 and 17.7, respectively. The Frontal Assessment Battery (FAB) had a mean score of 11.9 (SD = 2.9), indicating variable executive dysfunction. Language performance also varied, with mean scores of 11.3 on the Categorical Fluency Test (CFT–animal), 3.8 on the Verbal Fluency Test (VFT), and 42.3 on the Philadelphia Naming Test (PNT). Comprehension scores averaged 63.6 for sentence-level understanding and 4.6 for ADAS-Cog speech. Across the five imaging-derived regional volumes, the inferior frontal gyrus (mean = 9.0, SD = 1.6) and superior temporal gyrus (mean = 24.9, SD = 2.6) showed the highest values, while the pars opercularis (mean = 6.0, SD = 0.6) and insula (mean = 10.0, SD = 1.4) presented narrower ranges (Table 1).

**Table 1.** Summary of patient characteristics.

| Variable | Mean ± SD / N<br>(percentage) | Range (Min–Max) |
| --- | --- | --- |
| **Demographic Variables** |  |  |
| Age (years) | 63.59 ± 6.67 | 43 – 78 |
| Sex (Male) | 42 (48%) | - |
| Education (Higher) | 45 (52%) | - |
| **Behavioral Measures** |  |  |
| FBI–Negative Behaviors | 17.10 ± 7.61 | 3 – 36 |
| FBI–Disinhibited Behaviors | 17.72 ± 7.59 | 3 – 36 |
| FBI–Total | 34.70 ± 12.03 | 10 – 69 |
| Frontal Assessment Battery | 11.86 ± 2.91 | 6 – 18 |
| **Language and Cognitive Measures** |  |  |
| ADAS-Cog Speech | 4.62 ± 0.53 | 3 – 5 |
| Sentence Comprehension | 63.56 ± 12.53 | 46 – 86 |
| Categorical Fluency (Animals) | 11.31 ± 2.40 | 5 – 16 |
| Verbal Fluency (VFT–letters) | 3.76 ± 2.20 | 0 – 11 |
| Philadelphia Naming Test | 42.34 ± 5.20 | 28 – 50 |
| **Regional Brain Volumes** |  |  |
| Inferior Frontal Gyrus | 9.03 ± 1.55 | 7.4 – 11.9 |
| Pars Opercularis | 6.02 ± 0.59 | 5.4 – 7.0 |
| Insula | 10.01 ± 1.38 | 7.7 – 12.7 |
| Superior Temporal Gyrus | 24.85 ± 2.62 | 22.8 – 30.1 |
| Inferior Parietal Lobule | 17.07 ± 2.69 | 14.3 – 22.2 |

We first examined the pairwise associations between regional brain volumes and clinical performance using Spearman’s rank correlations across all 45 combinations of five imaging-derived predictors (inferior frontal gyrus, pars opercularis, insula, superior temporal gyrus, and inferior parietal lobule) and nine clinical outcome measures. The strongest unadjusted correlations emerged between verbal fluency tasks and regional atrophy. Specifically, inferior parietal lobule volume was negatively correlated with CFT-animal scores (ρ = –0.297, p = 0.0052), as were superior temporal gyrus (ρ = –0.294, p = 0.0057) and inferior frontal gyrus (ρ = –0.285, p = 0.0074). A similar trend was observed between the superior temporal gyrus and letter fluency (VFT) scores (ρ = –0.282, p = 0.0080). A positive correlation between insular volume and CFT-animal performance was also detected (ρ = 0.252, p = 0.0187). However, following Bonferroni correction for multiple comparisons (α = 0.0011), none of these associations remained statistically significant (smallest adjusted p = 0.233). No imaging– behavior correlation reached significance, either before or after correction, and all other correlations had ρ values below |0.25| (Figure 1).

**Figure 1.**
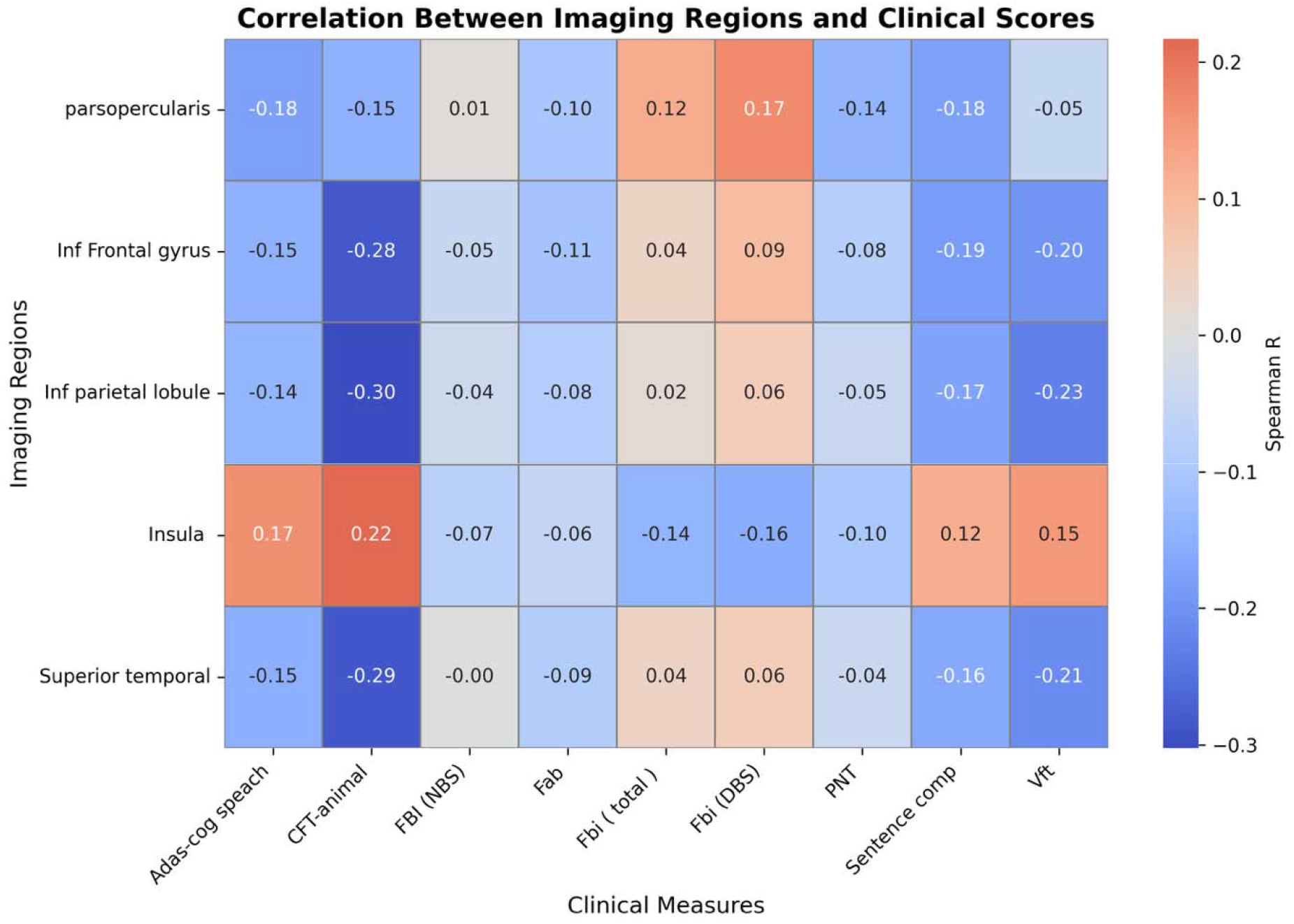
Spearman correlation heatmap

To investigate broader patterns, we aggregated the clinical measures into language-related (ADAS-Cog speech, sentence comprehension, CFT-animal, VFT, and PNT) and behavior-related (FBI-NBS, FBI-DBS, FBI-total, and FAB) domains. No significant associations were observed between any imaging region and behavior-related scores (all ρ < |0.20|, unadjusted p > 0.05), and none of the imaging–language correlations survived correction.

To reduce dimensionality, PCA was conducted on the five standardized regional volumes. Two components with eigenvalues >1.0 were retained, together accounting for 90.7% of the total variance (PC1 = 74.6%, PC2 = 16.1%). PC1 captured a global atrophy pattern, with strong positive loadings across all regions (range: 0.429–0.500), while PC2 was primarily driven by insular volume (loading = 0.961).

Multivariable logistic regression was performed to predict high behavioral impairment (FBI_total ≥ 30) using PC1, PC2, sex, age, and education as predictors. The model was based on 87 participants, yielding an events-per-variable (EPV) ratio of 11.4, exceeding the commonly accepted threshold of 10. The model converged without evidence of separation, and no predictor exhibited extreme coefficient estimates (all |β| < 10). Age was the only significant predictor (β = 0.107, SE = 0.041, z = 2.610, p = 0.009), corresponding to an odds ratio of 1.11 per year (95% CI: 1.03–1.21). PC1, PC2, sex, and education were not significant. The overall model fit was significantly better than the intercept-only null model (likelihood-ratio χ^2^, p = 0.0136).

To assess model discrimination, ROC analysis was conducted for three models: demographics-only (age and education), imaging-only (PC1 and PC2), and the full model. The AUC for the demographics-only model was 0.730, compared to 0.543 for imaging-only and 0.735 for the full model. At the Youden’s J threshold, the full model achieved a sensitivity of 0.65 and specificity of 0.80. Bootstrap resampling (n = 1000) revealed no significant improvement in AUC when comparing the full model to demographics alone (p = 0.453). Calibration plots indicated good alignment between observed and predicted probabilities, and Cook’s distances were uniformly < 0.09, suggesting no influential outliers.

Sex differences in regional brain volumes were assessed using parametric or non-parametric tests as appropriate, following checks for normality and variance homogeneity. After Bonferroni correction (α = 0.01), no significant differences were found in any region between males and females (all adjusted p > 0.05). Additionally, clinical scores were dichotomized at their respective medians and cross-tabulated with sex to explore potential associations. For ADAS-Cog speech, a nominal association was observed (χ^2^(1) = 4.13, p = 0.042), but this did not withstand Bonferroni correction (adjusted p = 0.380). No other clinical measure showed significant sex differences in distribution.

## Discussion

This study investigated the relationship between regional brain atrophy and clinical severity in patients with bvFTD. Although frontotemporal network dysfunction represents the canonical pathophysiological substrate of bvFTD, we found no significant associations between MRI-derived volumes of key frontotemporal regions and measures of disease severity after correction for multiple comparisons. Specifically, volumes of the inferior frontal gyrus, pars opercularis, insula, superior temporal gyrus, and inferior parietal lobule showed no statistically significant correlations with language performance (ADAS-Cog speech, sentence comprehension, category fluency, verbal fluency, picture naming) or behavioral impairment (FBI scales, FAB) following Bonferroni adjustment for 45 comparisons. Furthermore, principal components derived from these regional volumes failed to predict severe behavioral impairment (FBI ≥ 30) in multivariable logistic regression models, with age emerging as the sole significant predictor. These findings indicate that the relationship between regional brain atrophy and clinical phenotype in bvFTD may be more complex than previously understood.

The absence of significant correlations between regional volumes and language and behavioral measures after correction suggests a complex, nonlinear relationship between brain structure and function, rather than a true null association. Several interconnected mechanisms may explain this dissociation between macroscopic structural changes and functional decline. The focus on individual regional volumes may fundamentally overlook the more critical role of network-level dysfunction. Complex behaviors emerge from large-scale neural networks rather than isolated structures, with deficits often reflecting disruptions within corticolimbic or frontotemporal networks that single-node volumetric analysis cannot capture [12-15]. Furthermore, our investigation was limited to cortical regions of interest, yet subcortical structures are increasingly recognized as critical hubs in the networks underlying behavior. Previous work has directly correlated striatal atrophy with clinical measures of disease severity, suggesting these regions are key mediators of behavioral symptoms [16]. This is particularly relevant given that bvFTD, traditionally considered a frontotemporal cortical disease, exhibits growing evidence of significant subcortical involvement [17-19]. Specifically, the striatum may play a major role in generating behavioral and psychiatric symptoms through dysfunction of the limbic cortical-subcortical transmission circuits [19]. The dysfunction of this limbic circuit, with the limbic striatum serving as the key subcortical relay, has been implicated across multiple disease contexts as playing a major role in the pathogenesis of behavioral and psychiatric symptoms [19].

Second, the null findings at the group level may also reflect significant anatomical heterogeneity across participants. Different individuals can manifest similar clinical phenotypes through distinct patterns of neurodegeneration, with literature supporting at least four distinct anatomical subtypes in bvFTD, each with unique distributions of gray matter atrophy [20, 21]. Averaging across these distinct anatomical profiles in a single analysis could obscure significant relationships that exist only within specific, more homogeneous subgroups.

Third, our volumetric measures may lack the sensitivity to detect subtle but functionally critical microstructural changes. Diffusion tensor imaging can detect pathology before gray matter atrophy arises and correlates more directly with cognitive decline [22, 23]. Additionally, a temporal dissociation likely exists where neuropathological processes precede overt behavioral symptoms by months or years, creating a mismatch that cross-sectional designs cannot capture. The insidious onset of structural atrophy occurs long before patients reach clinical thresholds [24, 25]. Finally, the brain’s capacity for compensation may mask the functional impact of atrophy through cognitive or behavioral reserve. Neural plasticity allows for the recruitment of alternative networks to preserve function despite significant neuropathological burden. This compensatory mechanism may be particularly relevant in frontotemporal dementia, where enhanced connectivity between limbic cortical-subcortical circuits may initially reflect compensatory remodeling to maintain normal function as disease progresses [19, 26].

These findings collectively underscore that the relationship between brain structure and behavior in frontotemporal dementia is mediated by complex, dynamic processes involving network-level interactions, cortical-subcortical circuit dysfunction, anatomical heterogeneity, methodological limitations, temporal dissociation, and compensatory mechanisms, necessitating longitudinal designs and network-level analyses to fully elucidate these intricate relationships.

This interpretation is consistent with several recent studies that have documented similar challenges in establishing robust structure-function relationships in bvFTD using conventional grey matter volumetric measures. A recent multimodal neuroimaging study [27] demonstrated extensive cortical thinning and cortical mean diffusivity (cMD) increases across bilateral frontal and superior temporal regions, insula, and precuneus in bvFTD patients compared to controls. However, despite this widespread structural damage, cortical thickness in only a small cluster located in the right dorsomedial premotor cortex showed a significant relationship with disease severity as measured by the CDR plus NACC FTLD-SB scores, a finding that mirrors our own limited neuroanatomical-clinical correlations. Notably, this study also reported no significant relationships between verbal fluency test (VFT) scores and cortical thickness in bvFTD patients, consistent with our null findings for language measures. Interestingly, while naming ability (assessed via the Boston Naming Test) showed positive correlations with cortical thickness in medial and lateral temporal cortices and inferior parietal regions within the bvFTD subgroup specifically, areas that overlap with our superior temporal gyrus and inferior parietal lobule, these regional associations failed to reach significance in our more homogeneous sample, possibly reflecting differences in statistical power and sample heterogeneity. Importantly, the same study found that subcortical white matter measures (fractional anisotropy and mean diffusivity) and cortical mean diffusivity (cMD) revealed more extensive and significant clinical correlations than grey matter thickness, highlighting the potential limitations of structural volume-based approaches that we employed.

This pattern of attenuated structure-function relationships within homogeneous disease groups is further supported by recent longitudinal neuroimaging research [28] examining bvFTD, semantic variant primary progressive aphasia, and non-fluent agrammatic variant primary progressive aphasia. In a study examining 30 patients across the FTD spectrum (including 10 with bvFTD), significant associations were initially observed between baseline grey matter volume in bilateral frontal regions and cognitive decline trajectories using the Addenbrooke’s Cognitive Examination-Revised (ACE-R). However, when clinical diagnosis was included as a covariate in the statistical models, the predictive value of grey matter volumes was substantially weakened and no longer reached significance (P > 0.05), suggesting that structural measures primarily differentiate between disease categories rather than capturing meaningful within-group variance in clinical severity. This finding directly parallels our results, where structural volumes failed to predict behavioral impairment severity within our bvFTD cohort.

Similar methodological challenges have been reported in mixed-diagnostic studies examining neuropsychiatric symptoms and brain structure [29]. While this study demonstrated significant neuroanatomical-clinical correlations between behavioral symptoms (disinhibition, elation, aberrant motor behavior) and frontal volumes across a heterogeneous sample spanning mild cognitive impairment, Alzheimer’s disease, and bvFTD patients using the Neuropsychiatric Inventory (NPI), the authors noted that when diagnostic groups were analyzed separately, the associations were no longer statistically significant due to insufficient sample sizes within each subgroup. This observation suggests that the significant structure-function relationships observed in their combined analysis may have been driven primarily by between-group differences rather than within-group associations, similar to the pattern observed in our study and others [28]. The heterogeneous design of such studies, while maximizing variance in both neuropsychiatric symptoms and brain atrophy, may inadvertently inflate effect sizes by leveraging diagnostic group differences rather than revealing true neuroanatomical-clinical relationships within specific disease populations.

While this study provides valuable insights, several limitations should be considered when interpreting these findings. First, the cross-sectional design limits our ability to establish causal relationships and may not capture temporal dissociations where structural changes and functional impairment unfold at different disease stages. Longitudinal designs would provide valuable insights into the dynamic evolution of structure-function relationships across disease progression. Second, our sample size of 87 participants, while adequate for logistic regression, may have been insufficient to detect small to moderate correlations after multiple comparison correction. The relatively homogeneous nature of our bvFTD sample, while methodologically advantageous for reducing diagnostic confounding, may have resulted in range restriction of both structural and functional measures, making detection of correlational relationships more challenging. Third, our volumetric approach may have limited sensitivity to functionally significant changes. Gross volumetric measures cannot capture subtle microstructural alterations that advanced techniques such as diffusion tensor imaging [30, 31], resting-state functional connectivity [32], cortical thickness analysis [33], or PET biomarker studies [34] might detect [21]. These methods may be more sensitive to early pathological changes that precede volume loss and show stronger clinical associations. Fourth, disease heterogeneity represents a critical limitation, as bvFTD encompasses multiple underlying pathologies that may exhibit distinct structure-function relationships. Future studies would benefit from stratifying participants by C9orf72 repeat expansions, MAPT mutations, GRN mutations, and TDP-43 proteinopathy to examine structure-function relationships within specific pathological categories.

In conclusion, in this bvFTD cohort, structural volumes of key frontotemporal and perisylvian regions did not predict behavioral or language impairments after correction for multiple comparisons. These findings underscore the limitations of regional volumetric analysis in isolation and highlight the need for network-based, multimodal, and longitudinal approaches to better characterize structure–function relationships in bvFTD.

## Data Availability

All data produced in the present study are available upon reasonable request to the authors.

## Declaration

- Funding: This study was funded by Tehran University of Medical Sciences.
- Conflict of Interests/Competing Interests: Authors declare no conflict of interest.
- This study was approved by institutional review board of Tehran University of Medical Sciences.
- Written consent form was obtained from all participants.
- Data used in this study is accessible from corresponding author upon reasonable request.

## Notes

### Competing Interest Statement

The authors have declared no competing interest.

### Author Declarations

The study was approved by the ethics committee of Tehran University of Medical Sciences and adhered to the principles outlined in the Declaration of Helsinki. All participants or their legal representatives provided written informed consent in accordance with institutional ethics guidelines.

